# Prospective validation of smartphone-based heart rate and respiratory rate measurement algorithms

**DOI:** 10.1101/2021.03.08.21252408

**Authors:** Sean Bae, Silviu Borac, Yunus Emre, Jonathan Wang, Jiang Wu, Mehr Kashyap, Si-Hyuck Kang, Liwen Chen, Melissa Moran, John Cannon, Eric S. Teasley, Allen Chai, Yun Liu, Neal Wadhwa, Mike Krainin, Michael Rubinstein, Alejandra Maciel, Michael V. McConnell, Shwetak Patel, Greg S. Corrado, James A. Taylor, Jiening Zhan, Ming Jack Po

**Author notes:** Equal contribution, listed alphabetically by last name (Sean Bae, Silviu Borac, Yunus Emre, Jonathan Wang, Jiang Wu). Equal contribution (James A. Taylor, Jiening Zhan, Ming Jack Po).

## Abstract

Measuring vital signs plays a key role in both patient care and wellness, but can be challenging outside of medical settings due to the lack of specialized equipment. In this study, we prospectively evaluated smartphone camera-based techniques for measuring heart rate (HR) and respiratory rate (RR) for consumer wellness use. HR was measured by placing the finger over the rear-facing camera, while RR was measured via a video of the participants sitting still in front of the front-facing camera. In the HR study of 95 participants (with a protocol that included both measurements at rest and post exercise), the mean absolute percent error (MAPE) ± standard deviation of the measurement was 1.6% ± 4.3%, which was significantly lower than the pre-specified goal of 5%. No significant differences in the MAPE were present across colorimeter-measured skin-tone subgroups: 1.8% ± 4.5% for very light to intermediate, 1.3% ± 3.3% for tan and brown, and 1.8% ± 4.9% for dark. In the RR study of 50 participants, the mean absolute error (MAE) was 0.78 ± 0.61 breaths/min, which was significantly lower than the pre-specified goal of 3 breath/min. The MAE was low in both healthy participants (0.70 ± 0.67 breaths/min), and participants with chronic respiratory conditions (0.80 ± 0.60 breaths/min). Our results validate that smartphone camera-based techniques can accurately measure HR and RR across a range of pre-defined subgroups.

## Introduction

Measurement of heart rate (HR) and respiratory rate (RR), two of the four cardinal vital signs—HR, RR, body temperature, and blood pressure—is the starting point of physical assessment for both health and wellness. However, taking these standard measurements via a physical examination becomes challenging in telehealth, remote care, and consumer wellness settings.^1–3^ In particular, the recent COVID-19 pandemic has accelerated trends towards telehealth and remote triage, diagnosis, and monitoring.^4,5^ Although specialized devices are commercially available for consumers and have the potential to motivate healthy behaviors,^6^ their cost and relatively low adoption limit general usage.

On the other hand, with smartphone penetration exceeding 40% globally and 80% in the US,^7^ up to 3.8 billion individuals already have access to a myriad of sensors and hardware (video cameras with flash, accelerometers, gyroscope, etc) that are changing the way people interact with each other, and their environments. A combination of these same sensors together with novel computer algorithms can be used to measure vital signs via consumer-grade smartphones.^8–12^ Indeed, several such mobile applications (“apps”) are available, some with hundreds of thousands of installs.^13^ However, these apps seldom undergo rigorous clinical validation for accuracy and generalizability. Only a limited number of apps have undergone clinical evaluation for HR measurement (and/or atrial fibrillation detection),^14^ and the authors are unaware of any RR measurement apps that underwent clinical validation.

In this work, we present and validate two algorithms that make use of smartphone cameras for vital sign measurements. The first algorithm leverages photoplethysmography (PPG) acquired using smartphone cameras for HR measurement. PPG signals are recorded by placing a finger over the camera lens, and the color changes captured in the video are used to determine the oscillation of blood volume after each heart beat.^15^ In the second algorithm, we leverage upper-torso videos obtained via the front-facing smartphone camera to track the physical motion of breathing to measure RR. Herein, we describe both details of the algorithms themselves, in addition to reporting the performance of these two algorithms in prospective clinical validation studies. The studies sought to demonstrate reliable and consistent accuracy on diverse populations (in terms of objectively-measured skin tones, ranging from very light to dark skin) for HR and health status (with and without chronic pulmonary conditions) for RR.

## Methods

We conducted two separate prospective studies to validate the performance of smartphone-based HR and RR measurements (Figure 1). The user interfaces of the two custom research apps are shown in Supplementary Figure 1. The HR algorithm used PPG signals measured from the study participants placing their finger over the rear camera, and the enrollment for the corresponding validation study was stratified to ensure diversity across skin tones. The RR algorithm used video captures of the face and upper torso, and the enrollment for the corresponding validation study was stratified to capture participants with and without chronic respiratory conditions. The following sections detail each of the two studies.

**Figure 1.**
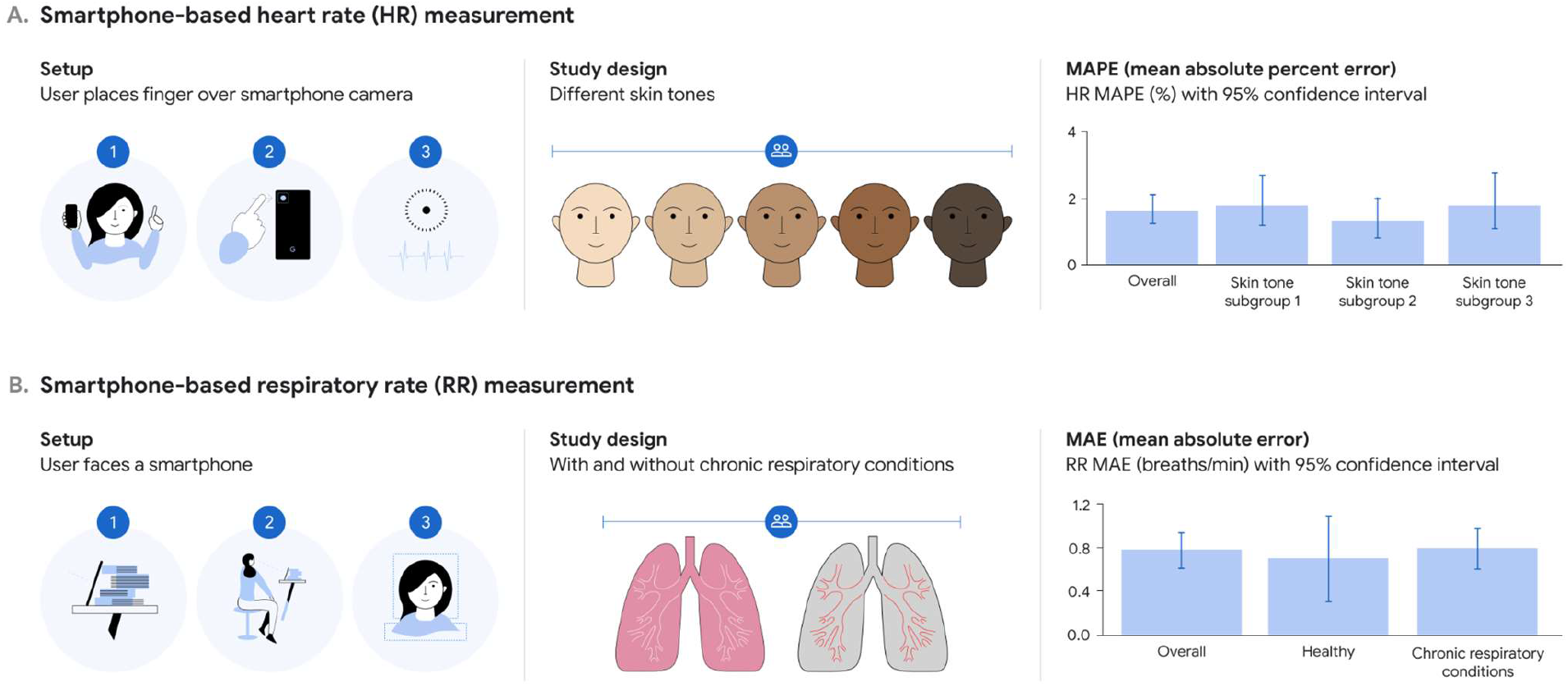
Smartphone-based monitoring of two key vital signs: (A) heart rate (HR) and (B) respiratory rate (RR). Left: the “setup” of how measurements are taken — with the finger over the rear-facing camera for HR, and using a video of the participant via the front-facing camera for RR. Middle: the study was designed to ensure generalization across skin tones for HR, and to participants with chronic respiratory conditions (chronic obstructive pulmonary disease and asthma) for RR. Skin-tone subgroup 1 corresponds to Fitzpatrick skin types 1–3 (very light, light and intermediate); subgroup 2 corresponds to types 4–5 (tan and brown); and subgroup 3 corresponds to type 6 (dark). Right: the main measurements were mean absolute percent error (MAPE) for HR, and mean absolute error (MAE) for RR.

### HR measurement

#### Algorithm description

Prior work in computer vision to extract heart rate from RGB (red-green-blue) video signals has leveraged manually extracted features in PPG signals from the finger for arrhythmia detection,^16^ ballistocardiographic movements from fingertips,^17^ red-channel PPG from fingertip videos,^18^ and the relationship between RGB channels.^19^

Our method estimates HR by optically measuring the PPG waveform from participants’ fingertips and then extracting the dominant frequency. First, several rectangular regions of interest (ROI) were manually selected from the video frames (linear RGB at 15 frames per second and at a resolution of 640×480 pixels). The chosen ROIs were the full frame, the left half, the right half, the top half, and the bottom half of the frames. Since camera pixels are illuminated non-homogeneously, signal strength can have spatial variations across pixels.^20^ Our method simultaneously analyzes different ROIs to identify one with the greatest signal-to-noise ratio (SNR).

Pixels in each ROI were averaged per-channel to reduce the effects of sensor and quantization noise, similar to prior work.^19^ The pulsatile blood volume changes were present as the AC components in these smoothed signals. We then weighted the three RGB waveforms to predict a single PPG waveform (weights 0.67, 0.33, and 0 for RGB respectively were empirically determined via grid search) for each ROI.

The resulting PPG waveforms were bandpass filtered to remove low- and high-frequency noise unlikely to be valid HR. Filter cut-off frequencies corresponded to a low of 30 beats/min and high of 360 beats/min. Next, large amplitude changes in PPGs due to motion were suppressed by limiting maximum allowed changes in amplitudes as 3x of moving average values. Then, frequency domain representations of PPGs were computed using the Fast Fourier Transform (FFT), from which we identified the dominant frequencies with maximum power. Because the PPG signals are periodic with multiple harmonics, the powers of the base frequencies were computed by summing the powers of their first, second, and third harmonics. SNRs were estimated for each ROI by computing the ratio between the power of the dominant frequency and the powers of non-dominant frequencies on a logarithmic scale. ROIs were filtered to only those with a SNR ≥0 dB, and the dominant frequency of the ROI with the highest SNR was reported. If no such ROI existed, no HR was reported.

#### Study design and participants

We performed a prospective observational clinical validation study to assess the accuracy of the study algorithm in estimating HR in individuals of diverse skin tones (Supplementary Figure 2A). Participants were enrolled at a clinical research site (Meridian, Savannah, GA) from October 2020 to December 2020. Study eligibility criteria were limited to excluding participants with significant tremor or inability to perform physical activity. The inclusion/exclusion criteria are detailed in Supplementary Table 1A. Study enrollment was stratified into 3 skin-tone subgroups (mapped to Fitzpatrick skin types;^21^ see Supplementary Table 2) to ensure broad representation: (1) types 1–3 (very light, light and intermediate); (2) types 4–5 (tan and brown), and (3) type 6 (dark). Skin tone was objectively measured from the participants’ cheek skin using a Pantone Capsure color matcher colorimeter (X-Rite, Grand Rapids, MI). Evidence suggests that darker skin tone is frequently under-represented in medical datasets,^22^ and that medical devices using optical sensors may be less accurate in those individuals.^23–25^ Therefore, the darkest skin-tone subgroup was intentionally oversampled to ensure the algorithm’s unbiased performance over various skin tones. Informed consent was obtained from all study participants in accordance with the tenets of the Declaration of Helsinki. The study protocol was approved by Advarra IRB (Columbia, MD; protocol no. Pro00046845). The clinical research site followed standard safety precautions for COVID-19 in accordance with the Centers for Disease Control and Prevention guidelines.

#### Data collection

Each participant underwent four 30-second data collection episodes with their index finger (of a hand of their choice) held directly over the study phone camera. Three of the 30-second episodes were collected at rest under various ambient brightness/lighting conditions: (1) with camera flash on and under regular ambient light, (2) with flash off and under regular ambient light, and (3) with flash off and under dim light. The fourth episode was collected post-exercise. In the original protocol, participants were instructed to ride a stationary bicycle for 30 seconds as strenuously as possible against light to medium resistance. After enrolling 37 participants, the exercise protocol was modified (with an IRB amendment) to achieve higher participant HR: participants were encouraged to achieve 75% of their maximal HR, which was calculated by subtracting the participant’s age from 220 beats/min. Exercise was completed either when the goal HR was achieved or when the participant asked to stop. The data were collected with flash off and under regular ambient light. Lighting conditions were controlled using 2 overhead and 1 front light emitting diode (LED) lights. The brightness level of the study environment was measured by a Lux meter (LT300 Light Meter, Extech, Nashua, NH) prior to each study. Measured brightness values were between 160 and 200 Lux for regular ambient light, and between 95 and 110 Lux for dim light.

The study was conducted using a mobile app deployed to a Pixel 3 smartphone running Android 10 (Google LLC, Mountain View, CA). HR estimation using the app was generally completed by the study participants following the in-app instructions, with the coordinators providing feedback on usage when needed. The reference HR was measured simultaneously during each data collection episode using a Masimo MightySat^®^ (Masimo, Irvine, CA), which is US Food and Drug Administration-cleared for fingertip measurement of pulse rate.^26^ The measurements were conducted in accordance with the vendor’s manual and taken at the end of each episode.

#### Statistical analysis

Each participant contributed up to 3 HR measurements at rest (with different lighting conditions), and up to 1 post-exercise. Measurements were paired observations: the algorithm-estimated HR and the reference HR from the pulse oximeter. For each algorithm measurement, up to three tries were allowed, and the number of tries required was recorded. The baseline characteristics (e.g., race, Fitzpatrick skin type, and skin-tone subgroup) of the participants who did not successfully complete algorithm estimation were compared with the rest of the study population using Fisher’s exact test. A paired measurement was dropped if either the algorithm estimation or reference measurement failed. The absolute error of each paired measurement was calculated as the absolute value of the difference between the algorithm-estimated and reference HR values. The mean absolute error (MAE) was the mean value of all absolute errors. Similarly, the absolute error from each paired measurement was divided by the reference value for that measurement and multiplied by 100 to produce the absolute percentage error. The mean absolute percentage error (MAPE) was the mean value for all absolute percent error values. The four (three at rest and one post-exercise; fewer if missing data) values from each participant were treated as statistically independent as the clustering effects (intra-participant correlation) were observed to be minimal.

The MAPE was the primary study performance criteria, as recommended by the current standards for HR monitoring devices.^27^ We also computed the standard deviation and 95^th^ percentiles. Sign tests were used to determine whether the absolute percentage errors were significantly <5%, both for the entire group of participants and the 3 skin-tone subgroups. Bland-Altman plots were used to visualize the agreement between the estimated values and the reference measurements and assess for any proportional bias (trends in the error with increasing values).^28^ The subgroup analysis across the three skin-tone subgroups was pre-specified.

#### Sample size calculation

HR data collection was planned for approximately 100 participants. Enrollment up to a maximum of 150 participants was allowed as we anticipated that some enrolled participants would be excluded prior to contributing HR data because they failed to meet the required skin tone distribution or because they were not able to exercise. Requirements for participant enrollment termination included ≥60 paired HR measurements in the dark skin tone subgroup and ≥20% of the post-exercise reference HR >100 beats/min. The study hypothesis was that MAPE was less than 5% in all of the 3 skin-tone subgroups. To estimate the sample size required for the study, we first conducted an IRB-approved feasibility study with a different set of 55 participants and similar measurements both at rest and post-exercise. In that study, the MAPE ± standard deviation was 0.91% ± 3.68%. Assuming double the mean and SD (i.e., 1.82% and 7.36%, respectively), a minimum of two paired measurements per participant, a skin-tone subgroup of ∼25 participants, and some dropout from incomplete data, the power to detect a MAPE > 5% was > 0.8.

### RR measurement

#### Algorithm description

Prior work in computer vision and sensors to extract RR from RGB video signals relied on changes in color intensities at specific anatomical points,^29,30^ tracking head motions,^31,32^ estimating optical flow along image gradients,^33^ or factorizing the vertical motion matrix.^34^

Our contactless method estimates RR by performing motion analysis in a ROI of the video stream that includes the base of the neck, shoulder line and upper torso of the participant. The main challenge was that variations in video due to respiratory motions are hard to distinguish from noise. We build on Eulerian, phase-based motion processing^35^ that is particularly suited for analyzing subtle motions. In each video frame, the position at each pixel was represented by the phase of spatially localized sinusoids in multiple scales (frequencies). To aggregate the information across scales and to obtain an intuitive representation of motion, we then transformed the spatial phases into optical flow by linearly approximating the position implied by each phase coefficient and averaging across scales. Using the Halide high-performance image library^36^, we were able to speed up the phase and optical flow computation to achieve real-time processing (1-4 ms per frame on Pixel 3a and Pixel 4 mobile devices).

Ensembling was then used to improve the predictive performance. A spectral-spatial ensemble was built in the following way. The respiratory ROI, together with the four quadrants obtained by equally subdividing the ROI defined five regions over which the vertical component of the optical flow was averaged. This resulted in five respiratory waveforms. Next, frequency-domain representations for each of these respiratory waveforms were computed via FFT, from which power spectra were computed. The power spectra were then aggregated to obtain a final ensembled power spectrum. Bandpass-filtering was performed to remove low and high frequencies unlikely to represent valid RRs. Filter cut-off frequencies corresponded to a low of 6 breaths/min and a high of 60 breaths/min. The maximum power frequency and the corresponding SNR value were computed from the ensembled power spectrum. The waveform corresponding to the entire ROI is used for displaying the breathing pattern to the user in the mobile app.

Often there was insufficient periodicity in the respiratory waveform (e.g., the participant briefly held their breath or changed their respiratory rate within the time window used for analyzing the waveform). To increase the robustness of RR estimation, the algorithm falls back on a time domain estimation method based on counting zero crossings of the waveform corresponding to the entire ROI whenever the SNR obtained via the FFT-based method was lower than a certain threshold. We tested two versions of the algorithm, differing only in terms of this threshold: SNR < −6.0 dB (“version A”) and SNR < −4.0 dB (“version B”). The higher value for the threshold in version B invoked the time domain estimation method more often, which was hypothesized to improve accuracy by improving robustness to irregular breathing.

#### Study design and participants

We performed a prospective observational clinical validation study to assess the accuracy of the study algorithm in measuring the RR in healthy adults and patients with chronic respiratory conditions (Supplementary Figure 2B). Participants were enrolled at a clinical research site (Artemis, San Diego, CA) between June 2020 and July 2020. Chronic respiratory conditions included moderate or severe chronic obstructive pulmonary disease (COPD) and asthma that was not well-controlled based on specific study criteria (Supplementary Table 1B). Also, participants with significant tremor were excluded. Further details and criteria are presented in Supplementary Table 1B. Informed consent was obtained from all study participants in accordance with the tenets of the Declaration of Helsinki. The study protocol was approved by Aspire IRB (now WCG IRB, Puyallup, WA; protocol no. 20201594). The clinical research site followed standard safety precautions for COVID-19 in accordance with the Centers for Disease Control and Prevention guidelines.

#### Data collection

Each participant underwent 30 seconds of data collection using a Pixel 4 smartphone running Android 10 (Google LLC, Mountain View, CA). The two algorithm versions (A and B) were tested sequentially. The participants followed the study protocol via instructions from the study app, without intervention from the study staff. Participants were prompted to prop the study phone on a table using provided common household items, such that the upper body was centered in the video capture (Figure 1). There were no specific requirements on the type of clothing worn during the study or additional custom lighting equipment. The in-app instructions guided the participants to wait several minutes after any active movement and to stay comfortable and breathe normally during the measurements.

During the data collection, RR was manually counted and recorded by two research coordinators. The two observers counted the number of breaths independently and blinded to the algorithm-estimated results. The agreement between the two measurements was high (Pearson correlation coefficient: 0.962; mean difference: 0.48 ± 0.88 breaths/min; range, 0–4). The mean of the two human-measured RRs, rounded off to the nearest integer, was taken to be the reference RR.

#### Statistical analysis and sample size calculations

Each participant contributed a single pair of measurements for each algorithm version, and the MAE was used as the primary evaluation metric. The study hypothesis was that MAE would be < 3 breaths/min. One-sample t-tests were done to determine whether the MAE was statistically significantly < 3 breaths/min. A pre-specified subgroup analysis was also performed, stratified by history of chronic respiratory conditions. In addition, *post-hoc* subgroup analyses were performed for age and race/ethnicity subgroups. Bland-Altman plots were used to analyze further for any trends in errors. Differences between the two algorithm versions were compared using a paired t-test.

To estimate the sample size required for the study, we first conducted an IRB-approved feasibility study with 80 healthy adults. Based on that MAE ± standard deviation (0.96 ± 0.72 breaths/min), a sample size of 50 participants was estimated to provide a power of > 0.99 to detect an MAE < 3. The power was also >0.99 for both the subgroup of 10 healthy participants and the subgroup of 40 with chronic respiratory conditions. If the MAE and standard deviation were doubled, the power would be >0.99, 0.71, and >0.99, respectively, for the full sample, healthy participants, and those with chronic respiratory conditions.

#### User experience survey

The participants were surveyed about their experience using the app. The questions covered their ease of setting up the phone at the desired angle to capture their face/torso; the clarity of the instructions; their comfort in using the app to assess their general wellness; their comfort in teaching someone else how to use the app; and their expected comfort in using the app several times a day (Supplementary Table 3).

## Results

### Heart rate measurement

A total of 101 participants were enrolled. After excluding one participant who was found to meet exclusion criteria (pregnancy), there were 100 valid enrollees. Among these, 3 were withdrawn due to skin tone distribution requirements, and 2 were withdrawn during data collection due to difficulty in data collection (such as inability to hold a phone properly or to obtain reference HR data). Thus, 95 participants completed data collection (Supplementary Figure 2). The participants had a mean age of 41.8 years, 75% were female, and skin-tone subgroups were evenly distributed as planned: 33% were subgroup 1 (very light, light and intermediate), 34% were subgroup 2 (tan and brown), and 34% were subgroup 3 (dark) (Table 1).

**Table 1.**
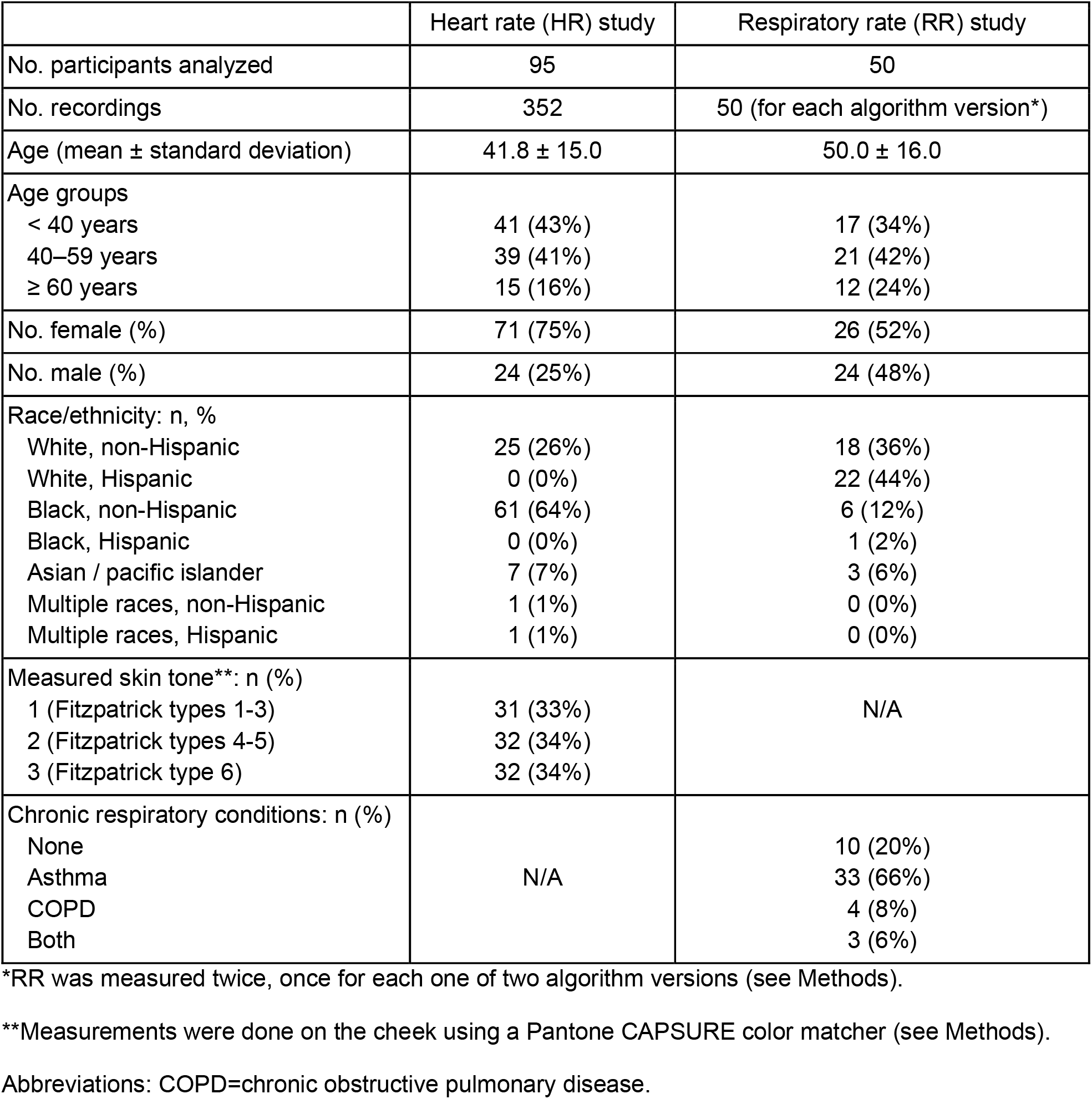
Baseline characteristics of the study participants.

From these participants, 379 total recordings were attempted. A valid HR was successfully obtained (see details on SNR in Methods) in 361 cases (95.3%). The success rate increased with retries up to 3 times: 316 measurements (83.4%) were successful on the first try, another 31 measurements (cumulative 91.6%) on the second try, and another 14 measurements (cumulative 95.3%) on the third try. The baseline characteristics of the 14 participants for whom HR values were not successfully reported by the study app for at least one measurement (due to low SNR) did not differ significantly from the remaining participants (Supplementary Table 4). In addition, a corresponding valid reference HR was not obtained for 9 recordings from 4 participants. The remaining 352 recordings with paired valid reference HR contributed to the final analysis (Supplementary Figure 2A). The average reference HR was 79.8 ± 14.6 beats/min overall, 75.5 ± 11.2 beats/min at rest, and 92.9 ± 16.6 beats/min post-exercise (Supplementary Table 5).

Compared to the reference HR, the MAPE of the overall study population was 1.63%, which was significantly lower than the pre-specified study target of 5% (p<0.001). The MAPE of 1.45% at rest and 2.39% post-exercise were also lower than the 5% target (p<0.001 for both). The MAPE showed a left-skewed distribution with a long tail (median, 1.14%; range, 0.0–50.6%). The MAPE by skin-tone subgroup was 1.77% for subgroup 1, 1.32% for subgroup 2, and 1.77% for subgroup 3, all of which met the study target of <5% (p<0.001 for all subgroups) (Figure 1 and Supplementary Table 5). We found no significant variation in MAPE across the three different lighting conditions.

Figure 2 shows the Bland-Altman plots for comparing the algorithm-estimated HR with the reference HR for the overall population and the three subgroups. Most observations (344/352, 97.8%) were within ±5 beats/min. Supplementary Figure 3 shows the Bland-Altman plots for HR stratified by at-rest versus post-exercise.

**Figure 2.**
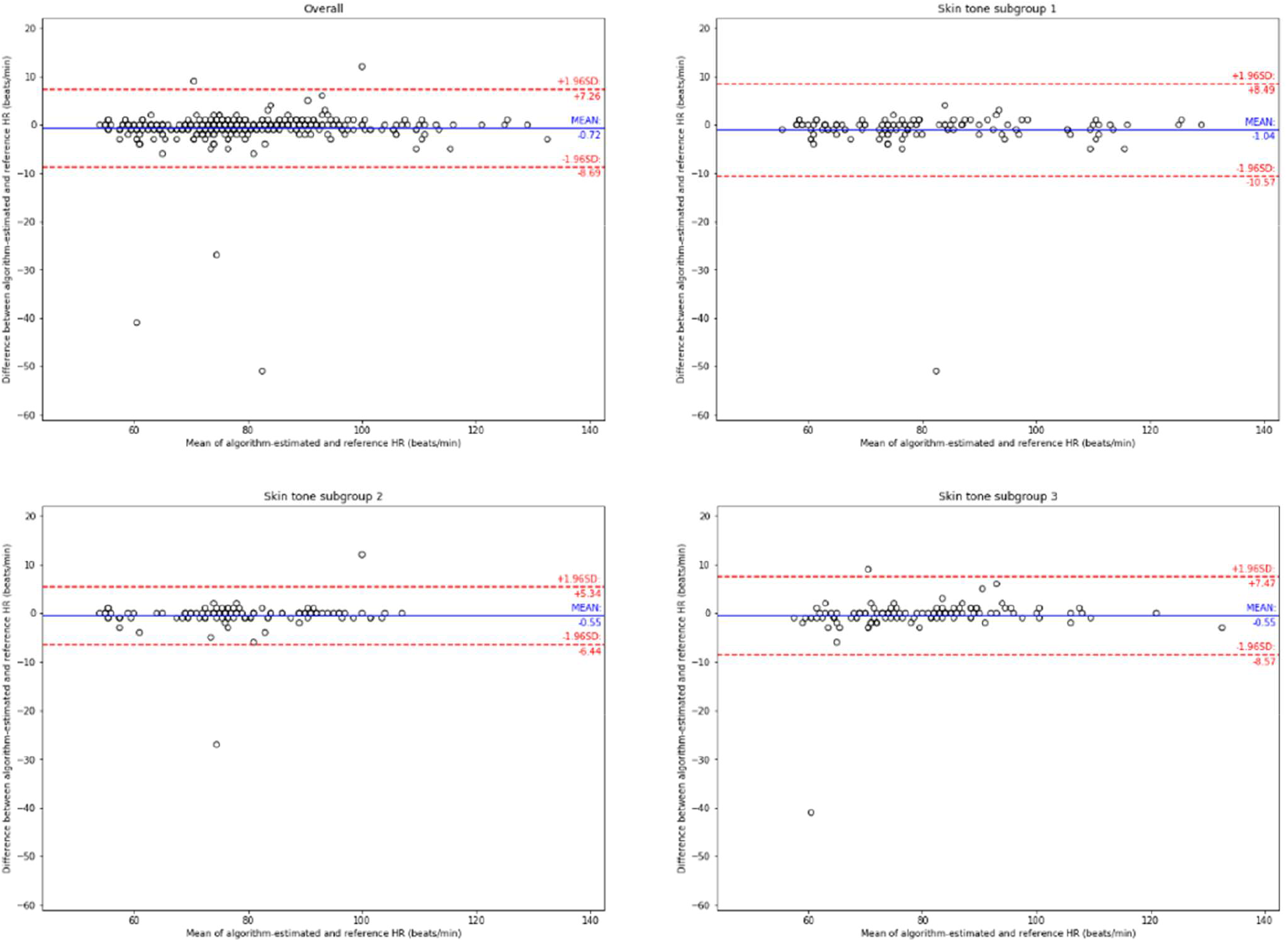
Bland-Altman plots for the heart rate (HR) study. Top to bottom and left to right: plots from the full study followed by subgroups based on skin type (see Figure 1). The reference HR was obtained from a pulse oximeter (see Methods). Dots represent individual participants; blue lines indicate the mean difference; red lines indicate the 95% limits of agreement (mean difference ± 1.96 standard deviations).

### Respiratory rate measurement

A total of 50 participants were enrolled in the RR study, including 10 healthy participants and 40 participants with chronic respiratory conditions (Supplementary Figure 2B). Self-identified baseline characteristics are presented in Table 1. The mean age was 50 years old; 80% were White, 14% were African American, and 46% were Hispanic. The average reference RR was 15.3 ± 3.7 breaths/min (Supplementary Table 6).

Both versions of the algorithm successfully estimated RR in all of the study subjects; thus, all of the 50 study participants contributed to the final analysis. The MAE in the overall study population was 0.84 ± 0.97 and 0.78 ± 0.61 breaths/min for algorithm versions A and B, respectively (Figure 1 and Supplementary Table 6), which were significantly lower than the pre-specified threshold of 3 breaths/min (p<0.001 for both). Each subgroup also showed MAE values significantly lower than the threshold: algorithm version A, 0.60 ± 0.52 breaths/min (p<0.001) for the healthy cohort and 0.90 ± 1.05 breaths/min (p<0.001) for the cohort with chronic respiratory conditions; algorithm version B, 0.70 ± 0.67 breaths/min (p<0.001) and 0.80 ± 0.60 breaths/min (p<0.001), respectively. No significant variations across age and race subgroups were seen (Supplementary Table 7).

Figure 3 shows the Bland-Altman plots for comparing the algorithm-estimated RR with the reference RR for the overall population and the two subgroups. All observations were within ±2 breaths/min of the reference RR for algorithm version B, while one observation was out of ±2 breaths/min for version A. The accuracy of the two algorithm versions did not differ significantly (p=0.70).

**Figure 3.**
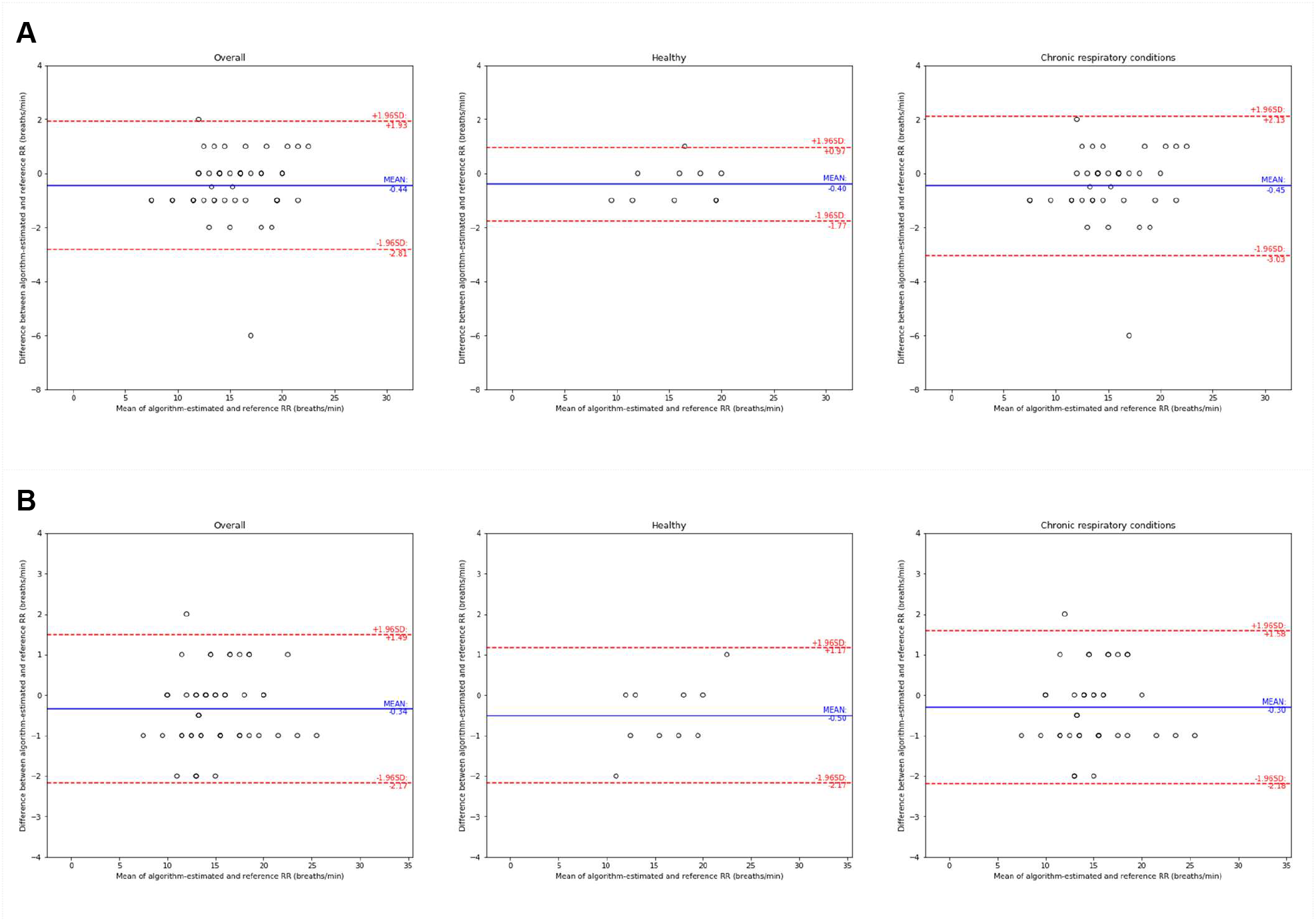
Bland-Altman plots for the respiratory rate (RR) study, for (A) algorithm version A and (B) algorithm version B. Left to right: plots from the full study followed by subgroups based on presence vs. absence of chronic respiratory conditions (see Figure 1). The reference RR was obtained by research coordinators manually counting breaths (see Methods). Dots represent individual participants; blue lines indicate the mean difference; red lines indicate the 95% limits of agreement (mean difference ± 1.96 standard deviations).

Supplementary Figure 4 shows the user experience survey results from the study participants. More than 90% of responses were positive, with participants reporting anticipated ease in setting up within a home environment, ease in following the instructions in the app, and comfort using the app to assess general wellness.

## Discussion

We report the results of two prospective clinical studies validating the performance of smartphone algorithms to estimate HR and RR. Both algorithms showed high accuracy compared to the reference standard vital sign measurements, with HR within 5% and RR within 3 breaths/min (the pre-specified targets). In addition, the HR estimation was robust across skin tones, and the RR estimation generalized to participants with common chronic respiratory conditions, COPD and asthma.

The accuracy of the HR algorithm is especially notable. A MAE less than 5 beats/min or a MAPE less than 10% are standard accuracy thresholds for HR monitors.^37,38^ The MAE of 1.32 beats/min in HR is lower than that reported for contemporary wearable devices (4.4 to 10.2 beats/min at rest), albeit with several differences in study design and population.^39^ The MAPE of 1.63% is comparable to the performance of current wearable devices. Shcherbina et al. tested six wrist worn devices to show a median error <5% for all across various activities and a median error of 2.0% for the best-performing device.^40^ Because skin tone can be a potential source of bias in medical devices,^23–25^ and the accuracy of PPG-based HR estimation can be affected by melanin’s light-absorbing property,^39,41^ we enrolled participants with diverse skin tones to validate the robustness of our HR estimation algorithm across skin tones.

For consumer-grade RR monitoring devices, there is no well-accepted accuracy standard.^27^ Our MAE of 0.78 breaths/min is comparable to that of professional healthcare devices, which have reported accuracy of ± 2–3 breaths/min.^42–45^ This could be a helpful reference point for future studies. In this study, we tested two algorithm versions for RR estimation that differed only in the SNR threshold. Our results suggest that this parameter had little impact on the accuracy or error rates.

This work supports the use of consumer-grade smartphones for measuring HR and RR. One application of these measurements is in fitness and wellness for the general consumer user. Specifically, an elevated resting HR or slower heart rate recovery after exercise has been linked to lower physical fitness and higher risk of all-cause mortality.^46,47^ Evidence suggests use of direct-to- consumer mobile health technologies may enhance positive lifestyle modification such as increased physical activity, more weight loss, and better diabetes control.^48–50^ Tracking one’s own health-related parameters over time by the general public can potentially increase motivation for a healthier lifestyle by providing an objective, quantifiable metric.^6^ Additionally, there exists strong evidence that regular physical activity is key to improving one’s health independent of age, sex, race, ethnicity, or current fitness level for maintaining cardiovascular health.^51^ Monitoring one’s HR is also an easy and effective way to assess and adjust exercise intensity, or enable smartphone-based measurement of cardiorespiratory fitness.^52–54^

With further clinical validation across broad populations, such smartphone-based measurement could also be useful in various settings, most notably telehealth where vital sign measurement is challenging due to the remote nature of the patient encounter.^55,56^ Though patients can in principle count their own HR or RR, this can be error prone due to factors such as biases that acute awareness of the self-examination can cause.^57,58^ Because the demand for remote triage, diagnosis and monitoring is burgeoning in the wake of the COVID-19 pandemic, there is increased attention being paid to accurate remote physical examination.^3,23,24^

There are several limitations in this work. First, our quantitative results focused on specific study devices (Pixel 3 and 4) and quantitative data on generalization to other devices will be needed. The current studies were also conducted in a controlled setting with structured study protocols. Though the participants used these features without significant study staff assistance, their ease of use in a general population will need further study. Next, our reference HR comes from a clinical PPG device instead of an electrocardiogram. There may be infrequent instances of electromechanical dissociation (such as various heart blocks, ventricular tachycardia, etc), in which a pulse rate measured at the periphery may not be the same with the reference electrical “heart” rate. Our HR algorithm is similarly subject to such errors because it relies on the pulsatile movement of blood in the fingertips. Further, awareness of self-measurement may affect users’ RR. A study demonstrated that people may have lower RR when they are aware that they are monitored by observers.^59^ It is yet unclear how awareness would affect RR measurements using apps in the absence of human-to-human interaction. In addition, though the study enrollment was optimized for diversity, this impacted the sample size in each subpopulation and the number of covariates that can be analyzed. Larger real-world validation that also controls for additional factors such as body-mass index will be helpful. Also, these clinical validation studies aimed to evaluate the algorithms at a “steady state” and further work will be needed to investigate acute clinical scenarios such as elevated or depressed HR or RR in urgent medical situations. Last but not least, although both HR and RR estimation algorithms met the predefined goals, observations with high deviation from the reference values were still produced by the algorithms (albeit infrequently). Future work to reduce errors is needed.

In addition to the clinical validation studies reported in this work, these HR and RR algorithms are currently undergoing additional broad usability testing across users, different Android devices, and different environments as we prepare to make the algorithms more widely accessible for consumers, beginning with Google Fit.

## Data Availability

Participants of this study did not consent to public sharing of their data, so the raw data are not available.

## Conclusion

We developed HR- and RR-measurement algorithms for smartphones, and conducted two clinical studies to validate their accuracy in various study populations. Both algorithm versions showed acceptable error ranges with a MAPE under 2% for HR and MAE under 1 breath/min for RR. These algorithms may prove useful in wellness settings such as fitness monitoring. Additional research is warranted before consideration for any future use in clinical settings, such as remote physical examination.

## Acknowledgements

This work would not have been possible without the contribution of many collaborators. We would like to thank Michael Righter and Shilpa Mydur for input on regulatory considerations, Xia Bellavia and Neil Smith for leading the mobile apps testing, Joe Nagle and Hyun Ji Bae for user interface / user experience (UI/UX) designs, Bhavna Daryani for data quality control in the heart rate validation study, Matt Shore for early stage product development, Rajroshan Sawhney for business development expertise, Benny Ayalew for help in data infrastructure, Justin Tansuwan and Rebecca Davies for visualization tools, Jassi Pannu, Tiffany Kang, and Jacinta Leyden for background research, and Robert Harle and Kapil Parakh for HR study protocol feedback. We are also grateful for the valuable manuscript and research feedback from Tiffany Guo, Jacqueline Shreibati, Ronnachai (Tiam) Jaroensri, Jameson Rogers, and Michael Howell. Lastly, we would like to thank Mark Malhotra, Bob MacDonald, Katherine Chou and (again) Shilpa Mydur and Michael Howell for their unwavering support.

## Author contributions

Y.E. and S.Bae developed the HR algorithms; S.Borac, J.Wu., N.W., M.Krainin, and M.R., developed the RR algorithms; J.Wang. and J.Wu, developed the mobile apps with the user interface for the clinical studies; J.Wang. developed software infrastructure for algorithms research; S.Bae. and S.Borac. developed software infrastructure for the HR study. E.S.T. provided data collection support for early-stage research; J.A.T., M.J.P., M.Kashyap, and J.C. developed the clinical study protocols; A.M. provided operational support for the clinical validation studies and managed inter-institutional communications; J.A.T. performed statistical analysis for RR study; J.A.T., S.Bae, and E.S.T. performed statistical analysis for the HR study; L.C. implemented the RR study and managed inter-institutional communications; L.C., M.Kashyap, and A.C. implemented the HR study and developed protocol optimizations; M.M. managed contracting and logistics required to begin the clinical validation studies; M.M. and A.C. performed quality control for the HR study data. S.K. and Y.L. provided research feedback and wrote the manuscript with the assistance and feedback of all authors. M.V.M. provided feedback on the HR study protocol; J.Z. oversaw technical developments and contributed to algorithm development; M.J.P. was the principal investigator for the clinical studies and developed the roadmaps encompassing the studies; S.P. and G.S.C. provided strategic guidance and funding for the project. All authors reviewed the manuscript.

## Competing Interests

All authors have completed the ICMJE uniform disclosure form at www.icmje.org/coi_disclosure.pdf and declare: all authors are employees of Google LLC except M.Kashyap and J.C. who are paid consultant of Google via Advanced Clinical; Google LLC may commercialize this technology or have patent rights now or in the future related to this paper. All the authors except M.Kashyap, S.K., and J.C. own Alphabet stock. M.J.P. is a member of the board of El Camino Health, advisory committee of ONC Interoperability Task Force, and scientific advisory board of National Library of Medicine, NIH.

## Funding/Support

This study was supported by Google Health.

## Role of the Funder/Sponsor

Google Health was involved in the design and conduct of the study; collection, management, analysis, and interpretation of the data; preparation, review, or approval of the manuscript; and decision to submit the manuscript for publication.

## Supplementary Information

### Supplementary Figures

**Supplementary Figure 1.**
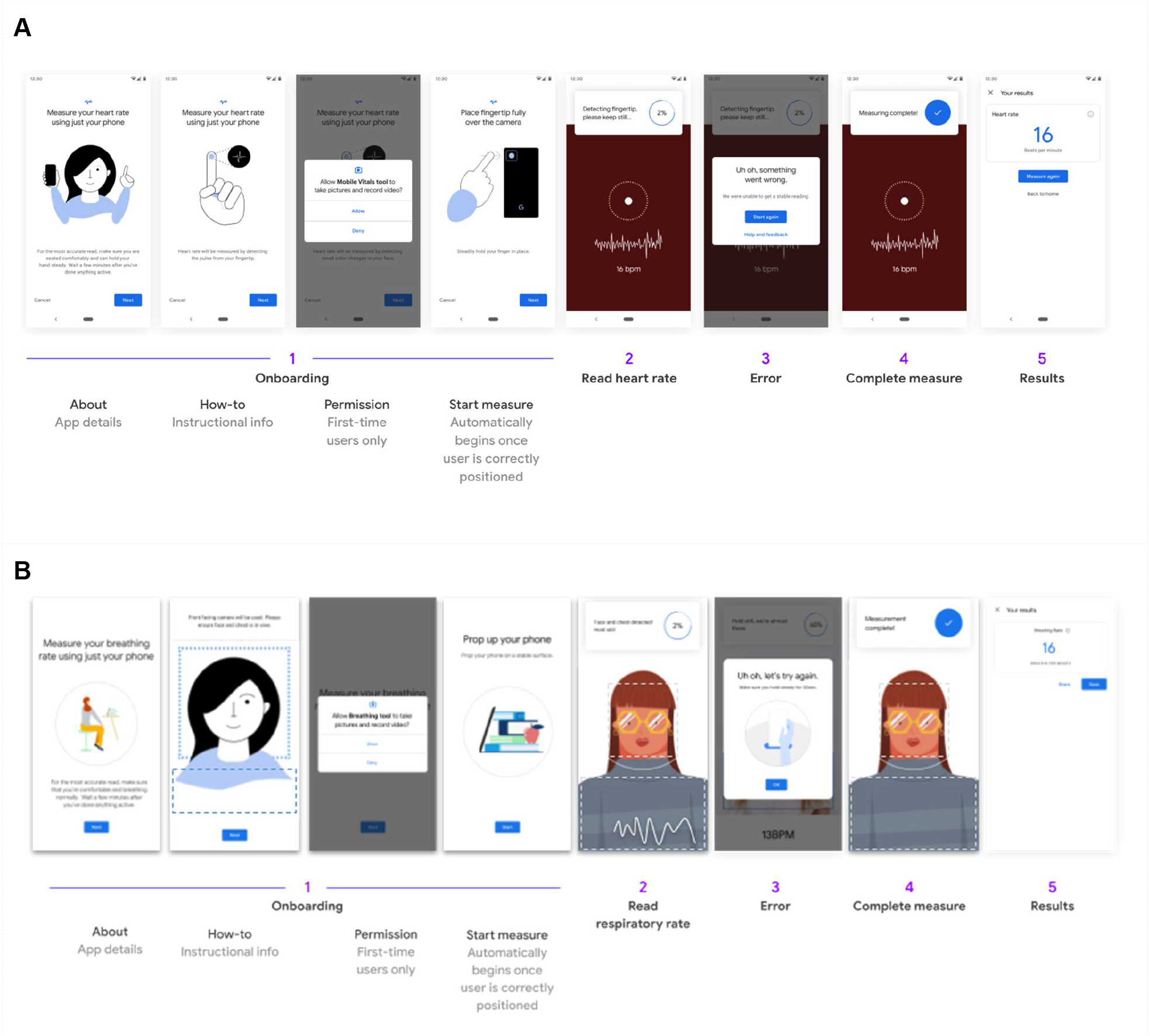
Detailed user interface for both heart rate (HR) and respiratory rate (RR) measurements.

**Supplementary Figure 2.**
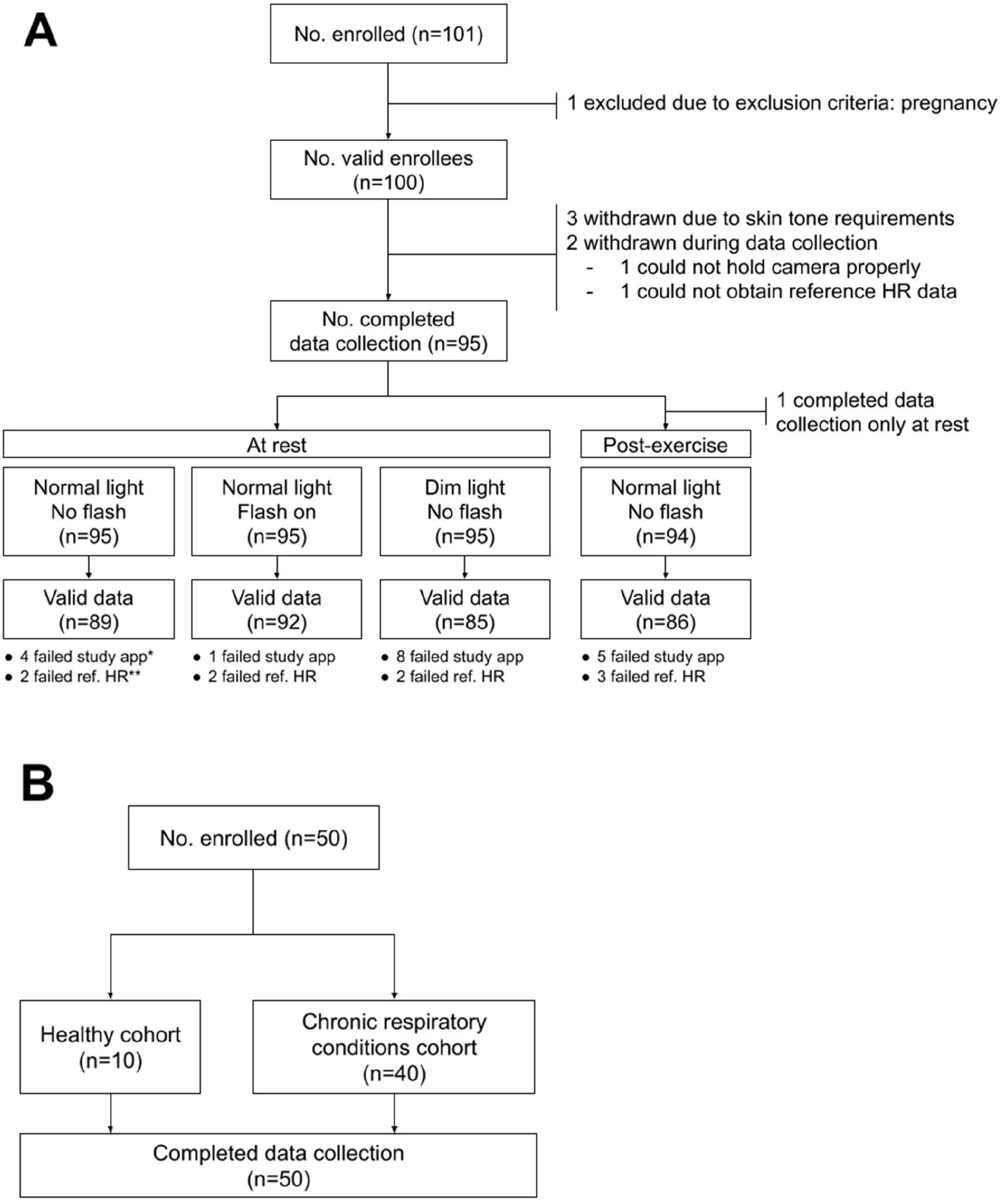
Number of participants enrolled and data analyzed in (A) the heart rate (HR) study and (B) the respiratory rate (RR) study. *Failed study application data collection due to signal-to-noise ratio <0; **Failed reference HR collection.

**Supplementary Figure 3.**
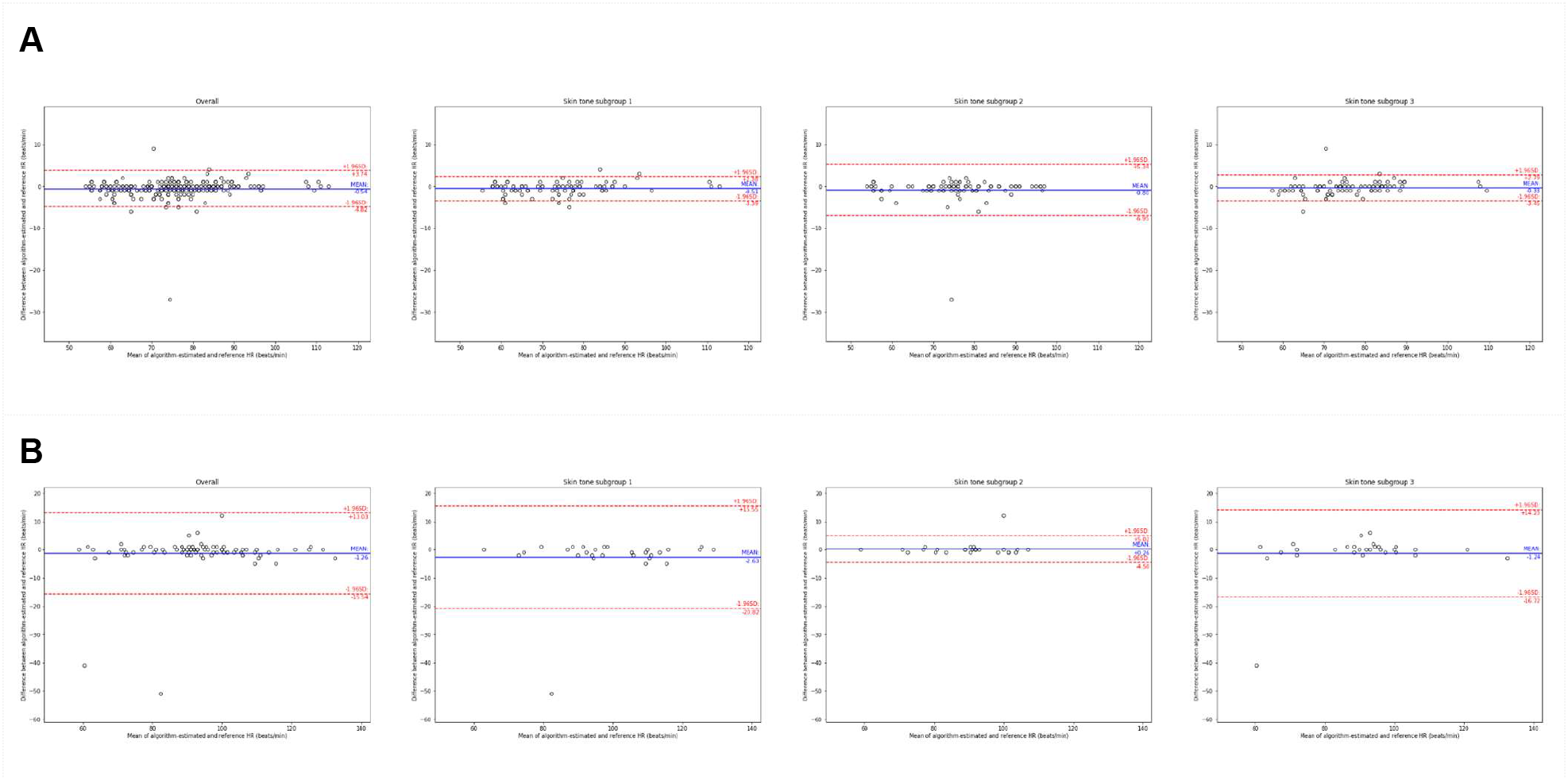
Additional Bland-Altman plots for the heart rate (HR) study (see Figure 2), for participants (A) at rest and (B) post-exercise. Left to right: plots from the full study followed by subgroups based on skin type (see Figure 1). The reference HR was obtained from a pulse oximeter (see Methods). Dots represent individual participants; blue lines indicate the mean difference; red lines indicate the 95% limits of agreement (mean difference ± 1.96 standard deviations).

**Supplementary Figure 4.**
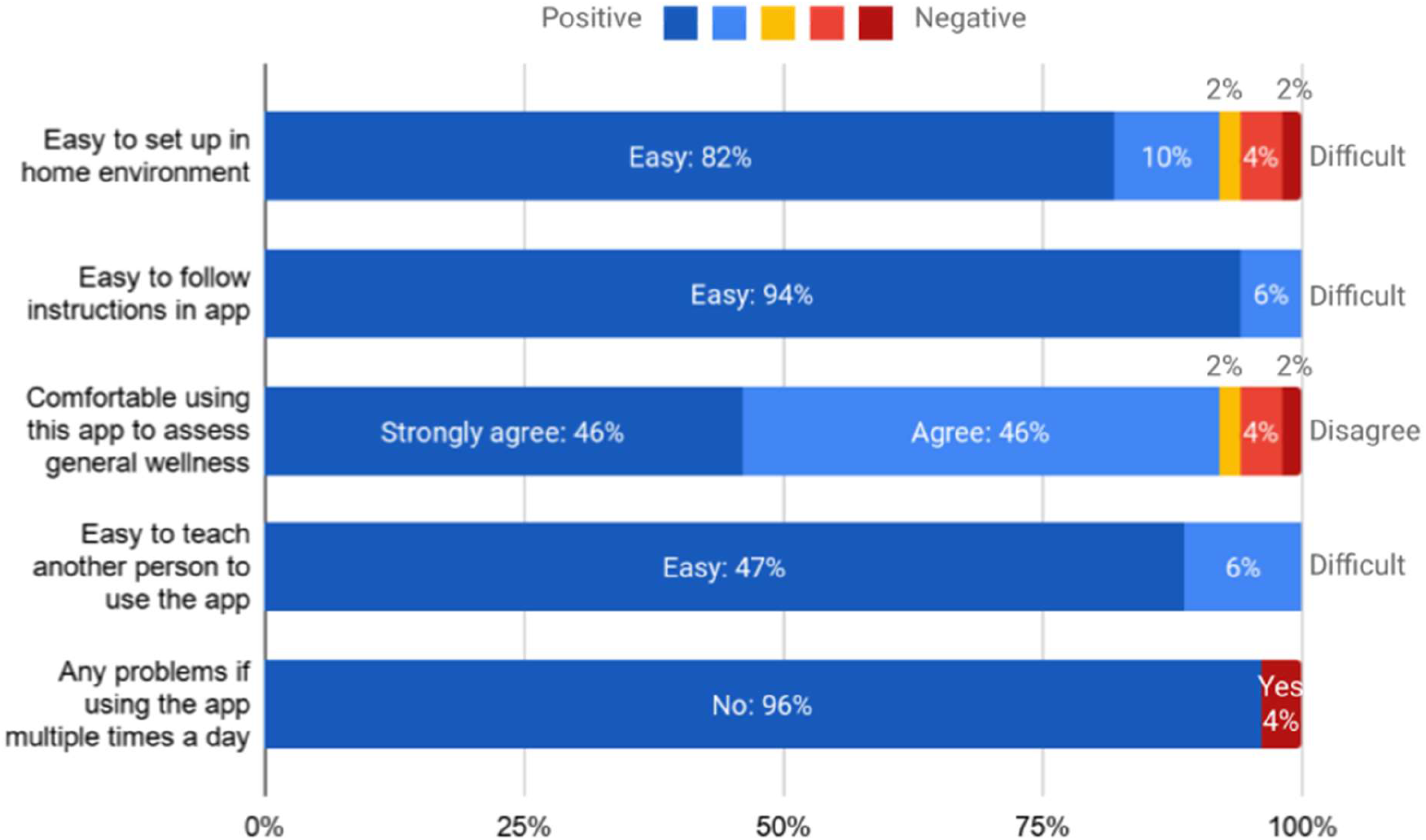
Participants’ survey results after the respiratory rate (RR) study. The exact survey questions are provided in Supplementary Table 3.

### Supplementary Tables

**Supplementary Table 1A.** Eligibility criteria of the heart rate (HR) study.

|  |  |
| --- | --- |
| **Inclusion criteria** | • Age > 18 years<br>• Participant is able and willing to provide written informed consent (Attached document: GH-VV-002 [ICF Heart Rate Study])<br>• Reads and speaks English. (Spanish speaking participants may be enrolled based on IRB requirements and/or availability of bilingual clinical study staff) |
| **Exclusion criteria** | • Unwilling or unable to remove makeup or face covering for study participation<br>• Pregnancy - Women who are, or believe they might be, pregnant<br>• Inability to understand study procedures and/or the informed consent process<br>• Other medical complications that preclude completion of the study as determined by the principal investigator at each study site<br>• Significant tremor that is present while seated at time of study as determined by the study coordinator (upper chest or face)<br>• Ineligibility for physical activity based on interpretation of PAR-Q by the site's principal investigator. Study participants who answer yes to one or more of the PAR-Q questions should be further assessed for eligibility by the site's principal investigator. |
| **Enrollment requirements** | **Age**<br>• ≥25% between 20–39 years old<br>• ≥25% between 40–59 years old<br>• ≥25% greater than 60 years old<br>**Skin Tones**<br>Measured by Pantone Device; see supplementary table 2 for details<br>• ≥30% Very light, light, intermediate (minimum 10 participants from each group, maximum 40 participants in total)<br>• ≥20% Tan, brown (minimum 10 participants from each group, maximum 30 participants in total)<br>• ≥30% Dark (minimum 30 participants) |

**Supplementary Table 1B.**
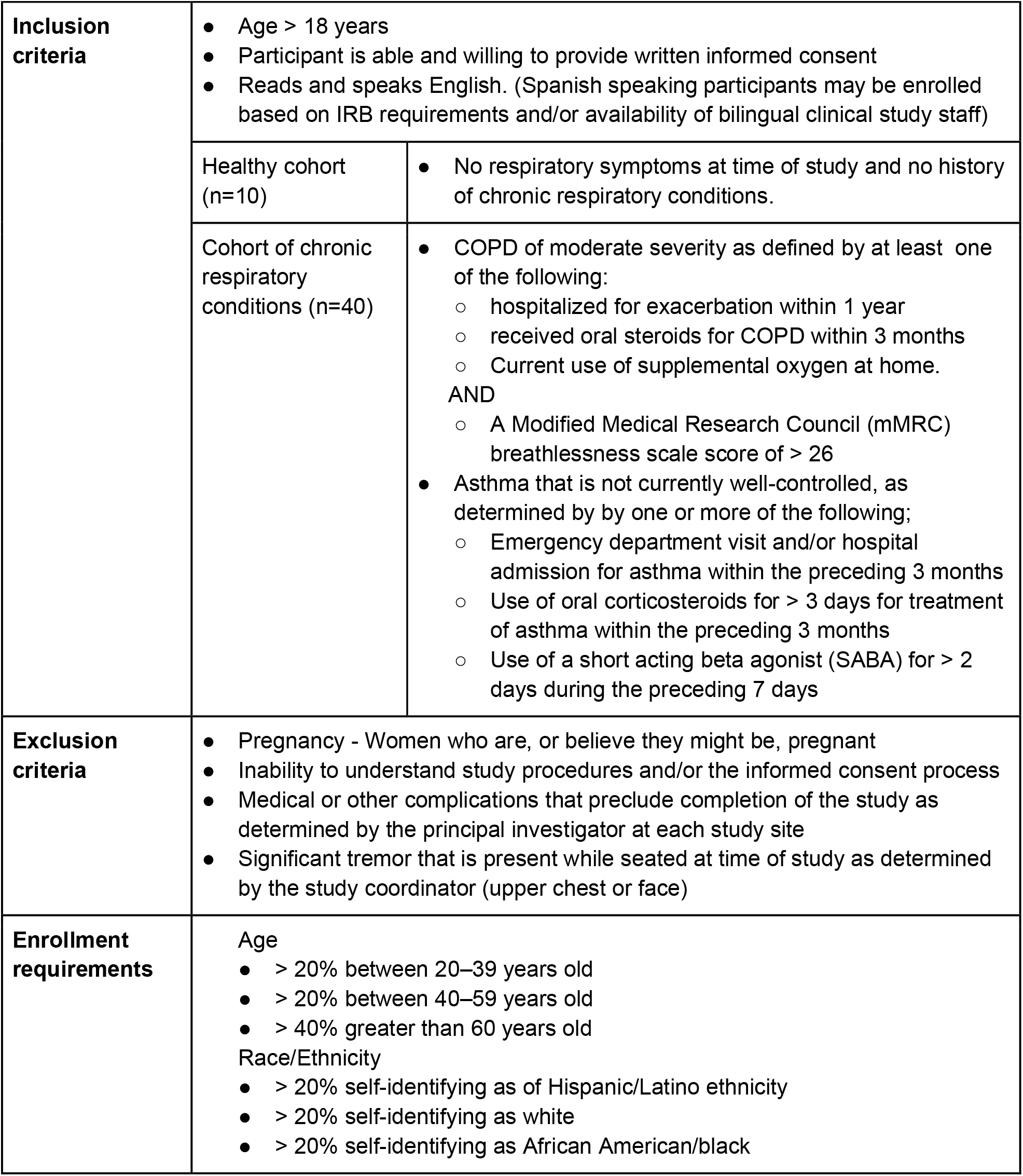
Eligibility criteria of the respiratory rate (RR) study.

**Supplementary Table 2.** Objective skin tone information was objectively measured by applying a Pantone CAPSURE color matcher to each participant’s cheek and mapping the parameters to a Fitzpatrick skin type. The numerical parameters were in the CIELAB color space, and the L* and b* parameters were converted to an individual typology angle (°ITA) as defined by the function [arctan(L* - 50)/b*] x 180/3.14159.

| $^{\circ}$ITA | Skin Tone Classification | Mapped to Fitzpatrick Skin Type |
| --- | --- | --- |
| $^{\circ}$ITA > 55° | Very Light | 1 |
| 41° < $^{\circ}$ITA < 55° | Light | 2 |
| 28° < $^{\circ}$ITA < 41° | Intermediate | 3 |
| 10° < $^{\circ}$ITA < 28° | Tan | 4 |
| -30° < $^{\circ}$ITA < 10° | Brown | 5 |
| $^{\circ}$ITA < -30° | Dark | 6 |

**Supplementary Table 3.** Participant experience survey on the respiratory rate (RR) measurement algorithm.

|  |  |  |  |  |  |
| --- | --- | --- | --- | --- | --- |
| How easy do you believe it would be to find a place to prop the phone up at home? | ☐<br>Easy | ☐<br>Somewhat Easy | ☐<br>Neither easy nor difficult | ☐<br>Somewhat difficult | ☐<br>Difficult |
| How would you rate the experience of following instructions in this application? | ☐<br>Easy | ☐<br>Somewhat Easy | ☐<br>Neither easy nor difficult | ☐<br>Somewhat difficult | ☐<br>Difficult |
| How much do you agree with the following statement, I would feel comfortable using this application to assess my general wellness? | ☐<br>Strongly agree | ☐<br>Agree | ☐<br>Neither agree nor disagree | ☐<br>Disagree | ☐<br>Strongly disagree |
| How well do you feel you could teach another person to use this application? | ☐<br>Easy | ☐<br>Somewhat Easy | ☐<br>Neither easy nor difficult | ☐<br>Somewhat difficult | ☐<br>Difficult |
| If you needed to use this app several times per day do you think there would be any problems? | ☐<br>Yes | ☐<br>No |  |  |  |
| (Optional) Do you have any other feedback you'd like to give on this application? | (Free description) |  |  |  |  |

**Supplementary Table 4.** Participant race and skin tone group stratified by signal-to-noise ratio (SNR) readings < 0 for the heart rate (HR) study.

|  | No. of participants with SNR<0 | No. of participants with SNR≥0 | P value* |
| --- | --- | --- | --- |
| Race/ethnicity |  |  |  |
| White, non-Hispanic | 5 (20.0%) | 20 (80.0%) | 0.798 |
| Black, non-Hispanic | 8 (13.1%) | 53 (86.9%) |  |
| Asian / pacific islander | 1 (14.3%) | 6 (85.7%) |  |
| Multiple races, non-Hispanic | 0 (0.0%) | 1 (100%) |  |
| Multiple races, Hispanic | 0 (0.0%) | 1 (100%) |  |
| Measured skin tone** |  |  |  |
| 1 (Fitzpatrick types 1-3) | 5 (16.1%) | 26 (83.9%) | 0.938 |
| 2 (Fitzpatrick types 4-5) | 4 (12.5%) | 28 (87.5%) |  |
| 3 (Fitzpatrick type 6) | 5 (15.6%) | 27 (84.4%) |  |
| Fitzpatrick type |  |  |  |
| 1 (Very light) | 0 (0.0%) | 1 (100%) | 0.194 |
| 2 (Light) | 1 (6.3%) | 15 (93.8%) |  |
| 3 (Intermediate) | 4 (28.6%) | 10 (71.4%) |  |
| 4 (Tan) | 2 (40.0%) | 3 (60.0%) |  |
| 5 (Brown) | 2 (7.4%) | 25 (92.6%) |  |
| 6 (Dark) | 5 (15.6%) | 27 (84.4%) |  |

\* P values were calculated by Fisher's exact test comparing participants with SNR<0 and those with SNR≥0
\*\* Fitzpatrick determination based on measurement with Pantone device and conversion to Fitzpatrick scale

**Supplementary Table 5.** Detailed results of the heart rate (HR) study.

| Session | Number of participants | Number of data points | Reference HR (beats/min) |  | MAE (beats/min) | MAPE (%) |  |  |
| --- | --- | --- | --- | --- | --- | --- | --- | --- |
| | | | Mean ± SD | Range | Mean ± SD | Mean ± SD | Median (IQR) | 95th percentile |
| Total population | 95 | 352 | 79.8 ± 14.6 | 54–134 | 1.32 ± 3.92 | 1.63 ± 4.27 | 1.14 (0.0–1.64) | 4.84 |
| At rest | 95 | 266 | 75.3 ± 11.1 | 54–113 | 1.02 ± 2.00 | 1.38 ± 2.47 | 1.20 (0.0–1.61) | 4.81 |
| Post-exercise | 94 | 86 | 93.9 ± 15.4 | 59–134 | 2.23 ± 7.05 | 2.39 ± 7.45 | 1.02 (0.0–1.75) | 5.42 |
| Subgroup 1: very light, light, and intermediate skin tone | 31 | 119 | 81.0 ± 17.4 | 56–129 | 1.53 ± 4.73 | 1.77 ± 4.46 | 1.27 (0.0–1.83) | 4.84 |
| At rest | 31 | 89 | 74.2 ± 11.8 | 56–113 | 1.02 ± 1.17 | 1.40 ± 1.60 | 1.28 (0.0–1.72) | 4.86 |
| Post-exercise | 31 | 30 | 101.0 ± 15.9 | 63–129 | 3.03 ± 9.15 | 2.90 ± 8.46 | 1.09 (0.82–2.09) | 4.36 |
| Subgroup 2: tan and brown skin tone | 32 | 113 | 78.8 ± 12.7 | 54–107 | 1.04 ± 2.87 | 1.32 ± 3.30 | 0.98 (0.0–1.37) | 4.86 |
| At rest | 32 | 86 | 75.4 ± 11.2 | 54–97 | 1.10 ± 3.04 | 1.44 ± 3.53 | 1.20 (0.0–1.43) | 4.99 |
| Post-exercise | 32 | 27 | 89.5 ± 11.3 | 59–107 | 0.85 ± 2.28 | 0.93 ± 2.43 | 0.0 (0.0–1.10) | 1.35 |
| Subgroup 3: dark skin tone | 32 | 120 | 79.7 ± 13.3 | 58–134 | 1.37 ± 3.89 | 1.77 ± 4.87 | 1.15 (0.0–1.64) | 4.48 |
| At rest | 32 | 91 | 76.3 ± 10.2 | 58–110 | 0.95 ± 1.32 | 1.31 ± 1.93 | 1.18 (0.0–1.56) | 3.94 |
| Post-exercise | 31 | 29 | 90.6 ± 15.9 | 61–134 | 2.69 ± 7.51 | 3.22 ± 9.27 | 1.12 (0.0–2.17) | 6.27 |

Abbreviations: HR, heart rate; MAE, mean absolute error; MAPE, mean absolute percentage error; SD, standard deviation; IQR, interquartile ranges

**Supplementary Table 6.** Detailed results of the respiratory rate (RR) study.

| Subgroups | N | Reference RR (breaths/min) |  | MAE (breaths/min) |  |  |
| --- | --- | --- | --- | --- | --- | --- |
| | | Mean ± SD | Range | Mean ± SD | Range | P value |
| Algorithm version A | 50 | 15.5 ± 3.6 | 8–22 | 0.84 ± 0.97 | 0–6 | <0.001 |
| Healthy | 10 | 16.0 ± 3.7 | 10–20 | 0.60 ± 0.52 | 0–1 | 0.001 |
| Chronic respiratory conditions | 40 | 15.4 ± 3.6 | 8–22 | 0.90 ± 1.05 | 0–6 | <0.001 |
| Algorithm version B | 50 | 15.3 ± 3.7 | 8–26 | 0.78 ± 0.61 | 0–2 | <0.001 |
| Healthy | 10 | 16.4 ± 3.7 | 12–22 | 0.70 ± 0.67 | 0–2 | <0.001 |
| Chronic respiratory conditions | 40 | 15.1 ± 3.7 | 8–26 | 0.80 ± 0.60 | 0–2 | 0.001 |

Abbreviations: RR, respiratory rate; MAE, mean absolute error; SD, standard deviation

**Supplementary Table 7.** Subgroup analysis for algorithm version B of the respiratory rate (RR) study.

| Subgroups | N | Reference RR (breaths/min) |  | MAE (breaths/min) |  |  |
| --- | --- | --- | --- | --- | --- | --- |
| | | Mean ± SD | Range | Mean ± SD | Range | P value |
| Age subgroups |  |  |  |  |  |  |
| <40 years old | 17 | 15.8 ± 4.4 | 10–26 | 0.79 ± 0.64 | 0–2 | <0.001 |
| 40–59 years old | 21 | 15.3 ± 3.1 | 10–22 | 0.79 ± 0.60 | 0–2 | <0.001 |
| $\geq 60$ years old | 12 | 14.8 ± 3.8 | 8–22 | 0.75 ± 0.62 | 0–2 | <0.001 |
| Race/ethnicity |  |  |  |  |  |  |
| White, non-Hispanic | 18 | 15.4 ± 4.1 | 8–26 | 0.72 ± 0.67 | 0–2 | <0.001 |
| Black, non-Hispanic | 6 | 18.0 ± 4.0 | 12–22 | 0.83 ± 0.41 | 0–1 | <0.001 |
| Hispanic/Latino ethnicity | 23 | 14.5 ± 2.6 | 10–19 | 0.83 ± 0.63 | 0–2 | <0.001 |
| Other | 3 | 16.3 ± 6.8 | 12–24 | 0.67 ± 0.58 | 0–1 | 0.010 |

## Notes

### Clinical Trial

This work was validation studies of a smartphone application for consumer wellness use.

### Funding Statement

This study was supported by Google Health. Google Health was involved in the design and conduct of the study; collection, management, analysis, and interpretation of the data; preparation, review, or approval of the manuscript; and decision to submit the manuscript for publication.

### Author Declarations

The HR and RR study protocols were approved by Advarra IRB (Columbia, MD; protocol no. Pro00046845) and Aspire IRB (now WCG IRB, Puyallup, WA; protocol no. 20201594), respectively.

## References

1. Tuckson RV, Edmunds M, Hodgkins ML. Telehealth. N Engl J Med. 2017;377(16):1585–1592.

2. Benziger CP, Huffman MD, Sweis RN, Stone NJ. The Telehealth Ten: A Guide for a Patient-Assisted Virtual Physical Examination. Am J Med. 2021;134(1):48–51.

3. Hyman P. The Disappearance of the Primary Care Physical Examination-Losing Touch. JAMA Intern Med. Published online August 24, 2020. doi:10.1001/jamainternmed.2020.3546

4. Hollander JE, Carr BG. Virtually Perfect? Telemedicine for Covid-19. N Engl J Med. 2020;382(18):1679–1681.

5. Cutler DM, Nikpay S, Huckman RS. The Business of Medicine in the Era of COVID-19. JAMA. 2020;323(20):2003–2004.

6. Rowland SP, Fitzgerald JE, Holme T, Powell J, McGregor A. What is the clinical value of mHealth for patients? NPJ Digit Med. 2020;3(1):4.

7. Contributors to Wikimedia projects. List of countries by smartphone penetration. Published April 29, 2014. Accessed January 12, 2021. https://en.wikipedia.org/wiki/List_of_countries_by_smartphone_penetration

8. Shao D, Liu C, Tsow F. Noncontact Physiological Measurement Using a Camera: A Technical Review and Future Directions. ACS Sensors. Published online 2020. doi:10.1021/acssensors.0c02042

9. De Ridder B, Van Rompaey B, Kampen JK, Haine S, Dilles T. Smartphone Apps Using Photoplethysmography for Heart Rate Monitoring: Meta-Analysis. JMIR Cardio. 2018;2(1):e4.

10. Sun Y, Thakor N. Photoplethysmography Revisited: From Contact to Noncontact, From Point to Imaging. IEEE Trans Biomed Eng. 2016;63(3):463–477.

11. Allen J. Photoplethysmography and its application in clinical physiological measurement. Physiol Meas. 2007;28(3):R1–R39.

12. Poh M-Z, McDuff DJ, Picard RW. Advancements in noncontact, multiparameter physiological measurements using a webcam. IEEE Trans Biomed Eng. 2011;58(1):7–11.

13. Koetsier J. The Top 10 Health & Fitness Apps Of 2020 Have One Thing In Common (Mostly). Forbes. Published October 5, 2020. Accessed January 20, 2021. https://www.forbes.com/sites/johnkoetsier/2020/10/05/the-top-10-health--fitness-apps-of-2020-have-one-thing-in-common-mostly/

14. O’Sullivan JW, Grigg S, Crawford W, et al. Accuracy of Smartphone Camera Applications for Detecting Atrial Fibrillation: A Systematic Review and Meta-analysis. JAMA Netw Open. 2020;3(4):e202064.

15. Hertzman AB. The Blood Supply of Various Skin Areas as Estimated by the Photoelectric Plethysmograph.; 1938.

16. Chong JW, Esa N, McManus DD, Chon KH. Arrhythmia discrimination using a smart phone. IEEE J Biomed Health Inform. 2015;19(3):815–824.

17. Zaman R, Cho CH, Hartmann-Vaccarezza K, Phan TN, Yoon G, Chong JW. Novel Fingertip Image-Based Heart Rate Detection Methods for a Smartphone. Sensors. 2017;17(2). doi:10.3390/s17020358

18. Chatterjee A, Prinz A. Image Analysis on Fingertip Video to Obtain PPG. Biomedical and Pharmacology Journal. 2018;11(4):1811–1827.

19. Wang W, den Brinker AC, Stuijk S, de Haan G. Algorithmic Principles of Remote PPG. IEEE Trans Biomed Eng. 2017;64(7):1479–1491.

20. Karlen W, Ansermino JM, Dumont GA, Scheffer C. Detection of the optimal region of interest for camera oximetry. Conf Proc IEEE Eng Med Biol Soc. 2013;2013:2263–2266.

21. Del Bino S, Bernerd F. Variations in skin colour and the biological consequences of ultraviolet radiation exposure. Br J Dermatol. 2013;169 Suppl 3:33–40.

22. Kinyanjui NM, Odonga T, Cintas C, et al. Estimating Skin Tone and Effects on Classification Performance in Dermatology Datasets. Published online October 29, 2019. Accessed February 17, 2021. http://arxiv.org/abs/1910.13268

23. Bickler PE, Feiner JR, Severinghaus JW. Effects of skin pigmentation on pulse oximeter accuracy at low saturation. Anesthesiology. 2005;102(4):715–719.

24. Sjoding MW, Dickson RP, Iwashyna TJ, Gay SE, Valley TS. Racial Bias in Pulse Oximetry Measurement. N Engl J Med. 2020;383(25):2477–2478.

25. Ries AL, Prewitt LM, Johnson JJ. Skin color and ear oximetry. Chest. 1989;96(2):287–290.

26. Bohnhorst B, Peter CS, Poets CF. Pulse oximeters’ reliability in detecting hypoxemia and bradycardia: comparison between a conventional and two new generation oximeters. Crit Care Med. 2000;28(5):1565–1568.

27. Consumer Technology Association: CTA Committees, Subcommittees and Working Groups. Accessed February 2, 2021. https://standards.cta.tech/kwspub/home/Committees/

28. Altman DG, Bland JM. Measurement in Medicine: The Analysis of Method Comparison Studies. The Statistician. 1983;32(3):307. doi:10.2307/2987937

29. Jorge J, Villarroel M, Chaichulee S, et al. Non-Contact Monitoring of Respiration in the Neonatal Intensive Care Unit. 2017 12th IEEE International Conference on Automatic Face & Gesture Recognition (FG 2017). Published online 2017. doi:10.1109/fg.2017.44

30. Massaroni C, Lopes DS, Lo Presti D, Schena E, Silvestri S. Contactless Monitoring of Breathing Patterns and Respiratory Rate at the Pit of the Neck: A Single Camera Approach. Journal of Sensors. 2018;2018:1–13. doi:10.1155/2018/4567213

31. Al-Naji A, Chahl J. Simultaneous Tracking of Cardiorespiratory Signals for Multiple Persons Using a Machine Vision System With Noise Artifact Removal. IEEE J Transl Eng Health Med. 2017;5:1900510.

32. Yang Q, Shen Y, Yang F, Zhang J, Xue W, Wen H. HealCam: Energy-efficient and privacy-preserving human vital cycles monitoring on camera-enabled smart devices. Computer Networks. 2018;138:192–200. doi:10.1016/j.comnet.2018.03.033

33. Chatterjee A, Prathosh AP, Praveena P. Real-time respiration rate measurement from thoracoabdominal movement with a consumer grade camera. Conf Proc IEEE Eng Med Biol Soc. 2016;2016:2708–2711.

34. Janssen R, Wang W, Moço A, de Haan G. Video-based respiration monitoring with automatic region of interest detection. Physiol Meas. 2016;37(1):100–114.

35. Wadhwa N, Rubinstein M, Durand F, Freeman WT. Riesz pyramids for fast phase-based video magnification. 2014 IEEE International Conference on Computational Photography (ICCP). Published online 2014. doi:10.1109/iccphot.2014.6831820

36. Ragan-Kelley J, Barnes C, Adams A, Paris S, Durand F, Amarasinghe S. Halide: a language and compiler for optimizing parallelism, locality, and recomputation in image processing pipelines. In: Proceedings of the 34th ACM SIGPLAN Conference on Programming Language Design and Implementation. PLDI ‘13. Association for Computing Machinery; 2013:519–530.

37. American National Standards Institute., Association for the Advancement of Medical Instrumentation. Cardiac Monitors, Heart Rate Meters, and Alarms. Arlington, Va. : Association for the Advancement of Medical Instrumentation, ©2002.; 2002.

38. CONSUMER TECHNOLOGY ASSOCIATION. Physical Activity Monitoring for Heart Rate (ANSI/CTA-2065). Accessed February 4, 2021. https://shop.cta.tech/products/physical-activity-monitoring-for-heart-rate

39. Bent B, Goldstein BA, Kibbe WA, Dunn JP. Investigating sources of inaccuracy in wearable optical heart rate sensors. NPJ Digit Med. 2020;3:18.

40. Shcherbina A, Mattsson CM, Waggott D, et al. Accuracy in Wrist-Worn, Sensor-Based Measurements of Heart Rate and Energy Expenditure in a Diverse Cohort. J Pers Med. 2017;7(2). doi:10.3390/jpm7020003

41. Fallow BA, Tarumi T, Tanaka H. Influence of skin type and wavelength on light wave reflectance. J Clin Monit Comput. 2013;27(3):313–317.

42. U.S. Food & Drug Administration. 510(k) Premarket Notification: Philips Biosensor BX100. Accessed February 25, 2021. https://www.accessdata.fda.gov/scripts/cdrh/cfdocs/cfpmn/pmn.cfm?ID=K192875

43. U.S. Food and Drug Administration. The C100 Contactless Breathing Monitor: 510(k) Premarket Notification. Accessed February 25, 2021. https://www.accessdata.fda.gov/scripts/cdrh/cfdocs/cfpmn/pmn.cfm?ID=K200445

44. MightySatTM Rx Fingertip Pulse Oximeter. Accessed February 2, 2021. https://techdocs.masimo.com/globalassets/techdocs/pdf/lab-10169a_master.pdf

45. Li T, Divatia S, McKittrick J, Moss J, Hijnen NM, Becker LB. A pilot study of respiratory rate derived from a wearable biosensor compared with capnography in emergency department patients. Open Access Emerg Med. 2019;11:103–108.

46. Jensen MT, Suadicani P, Hein HO, Gyntelberg F. Elevated resting heart rate, physical fitness and all-cause mortality: a 16-year follow-up in the Copenhagen Male Study. Heart. 2013;99(12):882–887.

47. Cole CR, Blackstone EH, Pashkow FJ, Snader CE, Lauer MS. Heart-rate recovery immediately after exercise as a predictor of mortality. N Engl J Med. 1999;341(18):1351–1357.

48. Flores Mateo G, Granado-Font E, Ferré-Grau C, Montaña-Carreras X. Mobile Phone Apps to Promote Weight Loss and Increase Physical Activity: A Systematic Review and Meta-Analysis. J Med Internet Res. 2015;17(11):e253.

49. Wu Y, Yao X, Vespasiani G, et al. Mobile App-Based Interventions to Support Diabetes Self-Management: A Systematic Review of Randomized Controlled Trials to Identify Functions Associated with Glycemic Efficacy. JMIR Mhealth Uhealth. 2017;5(3):e35.

50. Cadmus-Bertram LA, Marcus BH, Patterson RE, Parker BA, Morey BL. Randomized Trial of a Fitbit-Based Physical Activity Intervention for Women. Am J Prev Med. 2015;49(3):414–418.

51. U.S. Department of Health and Human Services. Physical Activity Guidelines for Americans, 2nd edition. Accessed January 15, 2021. https://health.gov/sites/default/files/2019-09/Physical_Activity_Guidelines_2nd_edition.pdf

52. Centers for Disease Control and Prevention. Target Heart Rate and Estimated Maximum Heart Rate. Centers for Disease Control and Prevention. Published October 14, 2020. Accessed January 15, 2021. https://www.cdc.gov/physicalactivity/basics/measuring/heartrate.htm

53. Arnett DK, Blumenthal RS, Albert MA, et al. 2019 ACC/AHA Guideline on the Primary Prevention of Cardiovascular Disease: A Report of the American College of Cardiology/American Heart Association Task Force on Clinical Practice Guidelines. Circulation. 2019;140(11):e596–e646.

54. Webster DE, Tummalacherla M, Higgins M, et al. Heart Snapshot: a broadly validated smartphone measure of VO2max for collection of real world data. bioRxiv. Published online July 4, 2020. doi:10.1101/2020.07.02.185314

55. Reilly BM. Physical examination in the care of medical inpatients: an observational study. Lancet. 2003;362(9390):1100–1105.

56. Verghese A, Charlton B, Kassirer JP, Ramsey M, Ioannidis JPA. Inadequacies of Physical Examination as a Cause of Medical Errors and Adverse Events: A Collection of Vignettes. Am J Med. 2015;128(12):1322-1324.e3.

57. Bickley LS. Bates’ Guide to Physical Examination and History Taking. Lippincott Raven; 2007.

58. Bickley L, Szilagyi PG. Bates’ Guide to Physical Examination and History-Taking. Lippincott Williams & Wilkins; 2012.

59. Hill A, Kelly E, Horswill MS, Watson MO. The effects of awareness and count duration on adult respiratory rate measurements: An experimental study. J Clin Nurs. 2018;27(3-4):546–554.

